# Operationalizing genomic epidemiology during the Nord-Kivu Ebola outbreak, Democratic Republic of the Congo

**DOI:** 10.1101/2020.06.08.20125567

**Authors:** Eddy Kinganda-Lusamaki, Allison Black, Daniel Mukadi, James Hadfield, Placide Mbala-Kingebeni, Catherine B Pratt, Amuri Aziza, Moussa M Diagne, Bailey White, Nella Bisento, Bibiche Nsunda, Marceline Akonga, Martin Faye, Ousmane Faye, Francois Edidi-Atani, Meris Matondo, Fabrice Mambu, Junior Bulabula, Nicholas Di Paola, Gustavo Palacios, Eric Delaporte, Amadou Alpha Sall, Martine Peeters, Michael R. Wiley, Steve Ahuka-Mundeke, Trevor Bedford, Jean-Jacques Muyembe Tamfum

## Abstract

The Democratic Republic of the Congo declared its tenth Ebola virus disease outbreak in July 2018, which has circulated primarily in the Nord Kivu province. In addition to standard epidemiologic surveillance and response efforts, the Institut National de Recherche Biomédicale implemented an end-to-end genomic surveillance system, including sequencing, bioinformatic analysis, and dissemination of genomic epidemiologic results to frontline public health workers. Here we report 538 new genomes from this outbreak; together with previously available sequence data (*n* = 48 genomes), this represents an unprecedented 17% of all laboratory-confirmed infections. To support outbreak response efforts, we reconstructed spatiotemporal transmission dynamics at broad and at fine scales as new data were available and disseminated the results via an interactive narrative-based platform. Our innovative system enables actionable information sharing between scientists and epidemiologists coordinating the day-to-day response on the time scales necessary to guide response efforts. The development of this genomic surveillance pipeline, within a resource-limited setting, represents significant technological and scientific progress in genomic epidemiology. Here we present a phylodynamic analysis of the outbreak as of February 2020, and describe the types of epidemiologic dynamics that we monitor the genomic data for, including resolution of co-circulating transmission chains, detection of superspreading events, inference of regions that act as transmission sources and sinks, and differentiation of closely linked cases versus propagated transmission. These findings have ameliorated the current outbreak response and are directly applicable to future outbreaks.

## Introduction

Since the first documented outbreak of Ebola virus disease (EVD) in Yambuku in 1976, outbreaks of EVD have occurred sporadically in the Democratic Republic of the Congo (DRC). The most recent outbreak began in late July 2018, when 26 cases of viral hemorrhagic fever were reported in Mabalako health zone, Nord Kivu province. Phylogenetic analysis of 48 Ebola virus (EBOV) genomes were used to distinguish the current outbreak from a previous one, which ended eight days before the Nord Kivu outbreak began ^1^. In June 2018, laboratory capacity to perform whole genome sequencing of EBOV had been established in-country, at the Institut National de Recherche Biomédicale (INRB) in Kinshasa. Sequencing capacity was extended to a field diagnostic lab in Butembo city, Katwa heath zone, in February, 2019. The establishment of sequencing labs in the DRC has allowed for continual genomic surveillance of this outbreak. At the time of writing, we had generated 586 full and partial genome sequences which were shared publicly shortly after sequencing at https://github.com/inrb-drc/ebola-nord-kivu. This represents ∼17% of known cases, a density of genomic surveillance that is both a significant achievement and a testament to the impact that building within-country capacity can have.

Comparative analysis of pathogen genomes can support traditional epidemiologic surveillance by improving our ability to detect and define clusters of related infections, facilitating detailed investigations of spatiotemporal dynamics, and monitoring for the emergence of genomic variants that infect or transmit more effectively. During the 2013-2016 West African EVD outbreak, analysis of viral genomic data was used to differentiate sexual EVD transmission from standard human-to-human transmission ^2^, to show that large, sustained case counts were attributable to many co-circulating transmission chains of varying sizes ^3^, and to detect the emergence of the A82V variant that rose to high frequency during the epidemic, most likely due to the variant’s increased infectivity in humans ^4,5^. For all of its utility, genomic surveillance also presents a challenge for public health agencies. Genomic surveillance generates large, complex datasets. Analyzing these data can require advanced computational infrastructure, as well as analysts trained in disciplines that have not historically been a part of public health, such as bioinformatics, computational biology, and data science ^6^. Presciently, Dudas and colleagues ^3^ postulated that as laboratory capacity for sequencing increased, genomic surveillance would become a routine part of outbreak response, but that analytic capacity would impede the timely release of genomic epidemiologic findings. Indeed, public health agencies’ ability to analyze and interpret genomic data within an epidemiologic context has lagged behind laboratory capacity to perform sequencing. This mismatch between the capacity to generate sequence data, and the capacity to interpret and communicate those inferences back to frontline public health workers, is a critical gap for the field to address.

During this outbreak we have used Nextstrain ^7^ to efficiently perform genomic epidemiologic analysis of sequence data as it is generated, and to share the results both publicly and privately through nextstrain.org. Beyond simply making the analyses available to public health officials, we have sought to increase their utility by improving how we communicate findings. To this end, we publish online, interactive situation reports describing our interpretation of the genomic data for the epidemiologists who are coordinating the day-to-day response. These situation reports, released in both English and French, are produced via Nextstrain Narratives ^8^, which allows simultaneous representation of genomic data visualizations and scientific interpretation.

In this paper we begin with an overview of the genomic surveillance system, describing sequencing intensity over the outbreak and patterns of data release. We then provide a phylodynamic analysis of the 586 publicly available genomes, discussing broad transmission dynamics between health zones over the course of the epidemic. Finally, we provide examples of the actionable, fine scale transmission dynamics we have monitored the genomic surveillance for. We describe the importance of these dynamics for frontline public health efforts such as contact tracing and infection control, and we discuss how we communicated our findings to public health workers.

### Online Methods

#### Ethics statement

Diagnostic specimens were collected as part of the DRC Ministry of Health public health emergency response; therefore, consent for sample collection was waived. All preparation of samples for sequencing, genomic analysis, and data analysis were performed on anonymized samples identifiable only by their laboratory or epidemiological identifier. USAMRIID and UNMC both determined that the generation of sequencing data for public health response was not research.

#### Sequence data generation

As described previously ^1^, clinical diagnostic specimens were collected from individuals presenting with EVD-like symptoms. Specimens were tested for the presence of EBOV RNA using the GeneXpert Ebola Assay (Cepheid, Sunnyvale, CA, USA). We sequenced a subset of all EBOV-positive samples; generally, samples are sequenced if they represent an epidemiologically important case, or if the case had an unusual contact history. Once samples were selected for sequencing, samples were sent to either the field genomics laboratory in Katwa or to INRB in Kinshasa. Samples were handled in a glovebox and RNA was extracted from the diagnostic specimen using the Viral RNA Mini kit (Qiagen). Samples were processed for sequencing using a hybrid capture method as described previously ^1^ or by an Amplicon based method ^9^. For hybrid capture sequencing, we used the KAPA RNA HyperPrep library preparation kit (KAPA Biosystems, Wilmington, MA, USA) with a spike-in of 20 ng HeLa RNA (Thermo Fisher, USA) and xGen Dual Index UMI Adapters (Integrated DNA Technologies, IA, USA) to prepare libraries. The libraries were enriched for EBOV using biotinylated probes (Twist Biosciences, USA) with the TruSeq Exome Enrichment kit (Illumina, San Diego, USA). For amplicon sequencing, the ThermoFisher 1st strand synthesis system was used to reverse transcribe RNA to cDNA. We amplified overlapping EBOV-specific amplicons according to a primer scheme generated from PrimalSeq ^9^ using Q5 DNA High-Fidelity Mastermix (New England Biolabs, Ipswich, MA) according to manufacturer’s specifications (primers are in table S1). Amplicons were quantified with the Qubit dsDNA High Sensitivity assay on the Qubit 4.0 instrument (Life Technologies, Carlsbad, CA) and then diluted to <500 ng for input into library preparation. Sequencing libraries were prepared using the Illumina Nextera DNA Flex kit (Illumina, San Diego, CA) with IDT for Illumina Unique Dual indexes. Libraries from both methods were quantified by qPCR with the KAPA Universal Library Quantification kit or by Qubit with the dsDNA High Sensitivity assay, andrun on an Illumina iSeq 100 or Miseq System for 2 × 150 cycles.

#### Bioinformatic and phylogenetic analysis

We used a custom bioinformatic pipeline to generate consensus genomes from the raw FASTQ-formatted sequencing output ^1,10^. De-identified metadata about the patient, diagnostic lab, and sequence quality were paired with the consensus genome. This additional data included the laboratory identifier of the sample, the epidemiologic identifier for the patient, the patient’s symptom onset date, the sample collection date, health zone, province, lab that performed the diagnostic testing, the sequencing date, and the percent genome coverage of the sequence. Phylogenetic analysis of all consensus genomes was performed using Nextstrain^7^, with updated builds occurring each time new sequences were released. Alignments were verified manually in Geneious (https://www.geneious.com/).

Our phylogenetic analysis pipeline utilises Augur version 6.3.0 (a component of Nextstrain), which performs a multiple sequence alignment with MAFFT v7.402^11^, computes a maximum likelihood phylogeny using IQ-TREE v1.6.6^12^, and temporally resolves this phylogeny using TreeTime v0.7.2 ^13^. We infer the health zone at internal nodes in the tree using the discrete trait reconstruction found in TreeTime. Resulting data are visualised using Auspice (a component of Nextstrain) which allows interactive exploration of the data.

#### Narrative construction and deployment

Upon release of new sequence data and completion of an updated Nextstrain build, we examined the phylogenies to determine where the new sequences clustered and to investigate epidemic dynamics apparent in the genomic data. Specific examples of these inferences are discussed in the Results. These inferences were written up in English and French as Nextstrain Narratives (Hadfield et al. 2019) and were disseminated through nextstrain.org. Briefly, Narratives are sourced from a Markdown file, which associates text with specific views of the data available on Nextstrain (see Supplemental Figure 1 for an example of an example). More information on writing, formatting, and deploying Narratives is available at https://nextstrain.github.io/auspice/narratives/how-to-write. The Narratives released to frontline public health workers contain sensitive patient information and thus are shared privately on a password-protected site. However, to illustrate what these situation reports are like, we have provided five narratives originally shared during September and October 2019, with this sensitive information redacted. Links to the online interactive versions and PDF copies of these narratives are available as Supplemental Information, and the data are available at https://github.com/blab/ebola-narrative-ms/ in branches “sitrep-2019-09-13”, “sitrep-2019-09-14”, “sitrep-2019-09-21”, “sitrep-2019-10-15” and “sitrep-2019-10-23”.

#### Data and code availability

All genomic surveillance data, including consensus genomes and de-identified metadata, are publicly available at https://github.com/inrb-drc/ebola-nord-kivu. Nextstrain Augur and Auspice are open-source and all source code can be found at https://github.com/nextstrain/augur and https://github.com/nextstrain/auspice All other analyses presented here are available as iPython notebooks at https://github.com/blab/ebola-narrative-ms/.

## Results

### Developing actionable genomic surveillance

Operationalizing genomic epidemiology in an outbreak setting requires consistent sequencing of samples in proportion to infection incidence, as well as rapid analysis and communication of results. To be actionable, return of results must occur quickly enough that the inferences can inform response efforts. As such, the success of genomic surveillance systems should be evaluated on the consistency and timeliness of sequencing, analysis, and data release.

As of February 2020, we have sequenced 586 EVD genomes from the Nord Kivu outbreak, 92% of which are included in a published analysis here for the first time. Samples have been sequenced over the full temporal span of the outbreak to date (Figure 1A). Despite the complex geographical and political situation in eastern DRC, sequencing intensity shows minimal geographic bias; the number of sequences from a health zone is proportional to the total number of cases reported within that health zone (Figure 1B). This lack of bias improves the validity of phylogeographic reconstructions of transmission dynamics.

**Figure 1:**
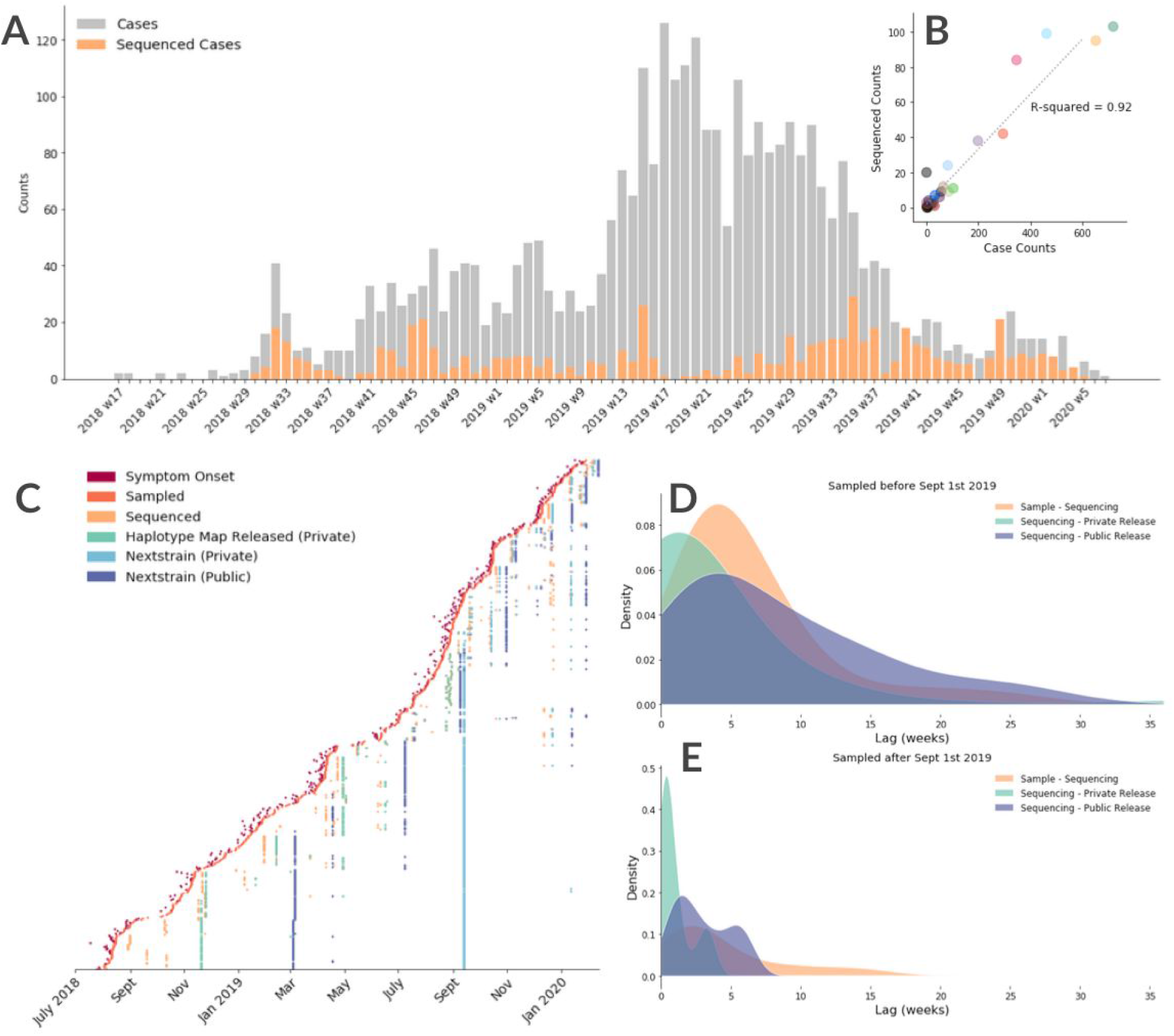
Progress of genomic surveillance over the course of the outbreak. (A) Total numbers of confirmed cases (grey) and sequenced cases (orange) by epi week; ∼17% of cases have been sequenced. (B) Correlation between the number of laboratory-confirmed cases reported in a health zone and the number sequenced cases from a health zone. (C) Time lags between sample collection and release of phylogenetic analyses. Three vertical lines, firstly for haplotype map release (teal), then for public Nextstrain release (purple), and then for private Nextstrain release (light blue) represent the initial roll-out of each of these platforms, when all sequence data up to that date were re-released on the new platform. The analysis in this paper uses the publicly released data. (D) Kernel density estimates of lag times between sample collection and sequencing (orange), between sequencing and private release of the data (teal), and between sequencing and public release of the data (purple), prior to September 2019. (E) Kernel density estimates of lag times between sample collection and sequencing (orange), between sequencing and private release of the data (teal), and between sequencing and public release of the data (purple), after switching to privately-released Nextstrain Narrative situation reports in September 2019.

To promote open data sharing and to facilitate potentially important insights from the international scientific and public health community, genomic data and de-identified metadata were released publicly through virological.org, GitHub, and Nextstrain as they were generated. As the genomic surveillance system developed over the outbreak, the time between sequencing and data release decreased (Figure 1C). While releasing data publicly supports open science in the broader community, data release alone does not necessarily make these data actionable. Rather, interpretations of the analyzed data must be circulated rapidly to be useful for public health response. Initially, we constructed and disseminated haplotype maps which were manually annotated with epidemiologic information. These visualizations were shared with the response team as PDFs, and were subsequently discussed at regular emergency operations meetings. In September 2019, we started to routinely run bioinformatic analysis in Kinshasa, and implemented private data sharing to accredited parties through nextstrain.org. This allowed us to transition from generating haplotype maps to constructing phylogenies with the Nextstrain pipeline, which improved the efficiency and scalability of analyses. Using Nextstrain enabled us to include genomes that were less than full length in the analyses as phylogenetic reconstruction methods can handle uncertainty at sites in an alignment. Secondly, we were able to generate temporally-resolved trees in addition to visualizations of genetic distance; this allowed estimation of when transmission chain movement occurred. Finally, the Nextstrain analytic pipeline enables automated annotation of many of the traits that we used to manually annotate in the haplotype maps. This reduced the average time between sequencing and sharing of phylogenetic reconstructions to 6.6 days (standard deviation 7.8 days). Public data sharing occured an average of 13.4 days later (Figure 1D,E).

In September 2019 we also began using Nextstrain Narratives ^8^ to deliver genomic epidemiological situation reports. Nextstrain Narratives provide simultaneous representation of genomic data visualizations alongside a narrative scientific interpretation. They are also interactive, providing users detailed information about the data on demand. This switch to using Narratives had various advantages; we could include more genomic data in the analyses and we could present both genetic divergence and temporally-resolved phylogenies. The Nextstrain Narrative situation reports, written in both English and French, were shared electronically as well as being presented to field epidemiologists at emergency operations centre meetings, typically held in Beni.

When circumstances have been ideal, we have performed diagnostic testing, sample transportation, and sample preparation for sequencing in as little as 4 days, with sequencing and data analysis taking an additional 2 to 3 days. Indeed, we have made genomic epidemiological inferences available to the response team in as little as 7 days after sample collection from a patient. However, the time between sample collection and sequencing is usually longer than this. On average, a sample is sequenced 41 days after collection from the patient, with a standard deviation 37 days. This relatively large lag results from logistical challenges and political instability, which we describe in greater detail in the Discussion.

### Broad-scale dynamics of EVD

From phylogeographic analysis of 586 publicly available EBOV genomes collected between July 2018 and January 2020, we describe broad patterns of the outbreak in space and time. The genomic data suggest that the outbreak began in late-July 2018 in the Mabalako health zone (Figure 2A), a finding that agrees with case surveillance data ^1^. There is only evidence for a single zoonotic spillover event, with all subsequent cases caused by human-to-human transmission. Transmission to the nearby health zones of Beni and Mandima appears to have occurred early after the outbreak started (Figure 2A,B). The genomic data suggest that there were multiple introductions of EVD from Mabalako into Beni (Figure 2A). One of these introductions, which we estimate occurred in August 2018 (95% CI: Aug 5, 2018 – Aug 18, 2018) led to extensive onward transmission into other health zones, becoming the primary circulating lineage in this outbreak (Figure 2A, “Primary Outbreak Clade”). We also observe movement of transmission chains back into previously affected health zones. For example, the primary outbreak clade moved from Beni into Kalunguta around the end of September 2018 (95%CI: Aug 19, 2018 – Oct 9, 2018), which later led to multiple introductions into Katwa during October 2018 through January 2019. One of the transmission chains circulating in Katwa then moved back into Beni some time between late-February and early-May 2019 (Figure 2A). A secondary, sustained lineage resulted from an introduction from Beni to Katwa sometime between August and October 2018 (Figure 2A, “Secondary Outbreak Clade”). This lineage later moved into Mandima, Rwampara, and back into Katwa. Although smaller than the primary outbreak clade, this secondary lineage has persisted throughout much of the outbreak, with descendents sampled as recently as September 2019.

**Figure 2:**
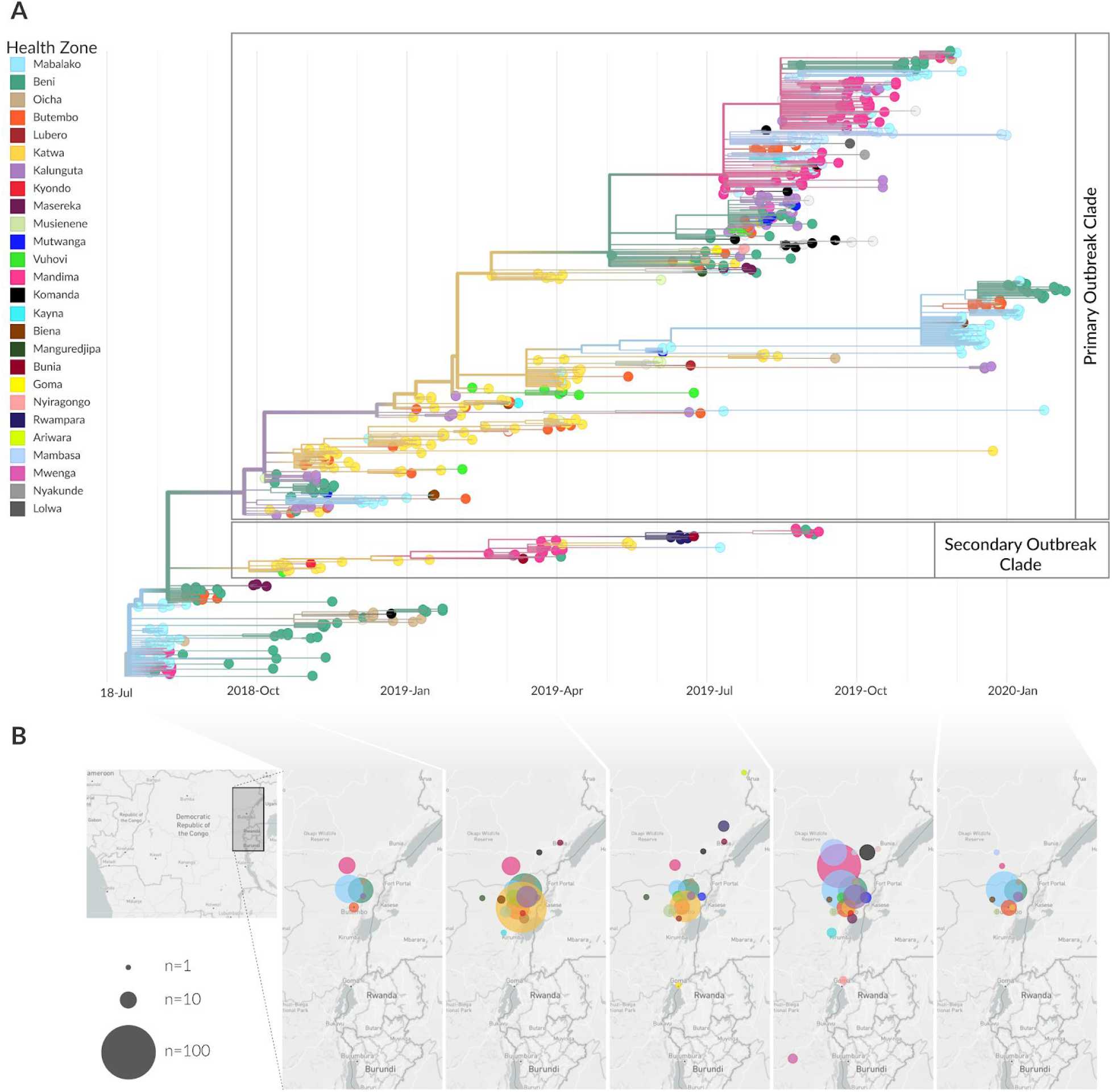
Broad scale spatiotemporal dynamics of the EVD outbreak in Nord Kivu province DRC. (A) temporal phylogenetic reconstruction of 586 genomes colored by reporting health zone, with clades of interest annotated; the health zone of internal branches is inferred via a discrete model and reduced confidence is conveyed by transitioning colors to gray. (B) Geographical spread of samples over five disjoint time intervals which span the entire outbreak (inferred transmissions removed for clarity). Figure adapted from Nextstrain visualizations.

**Figure 2:**
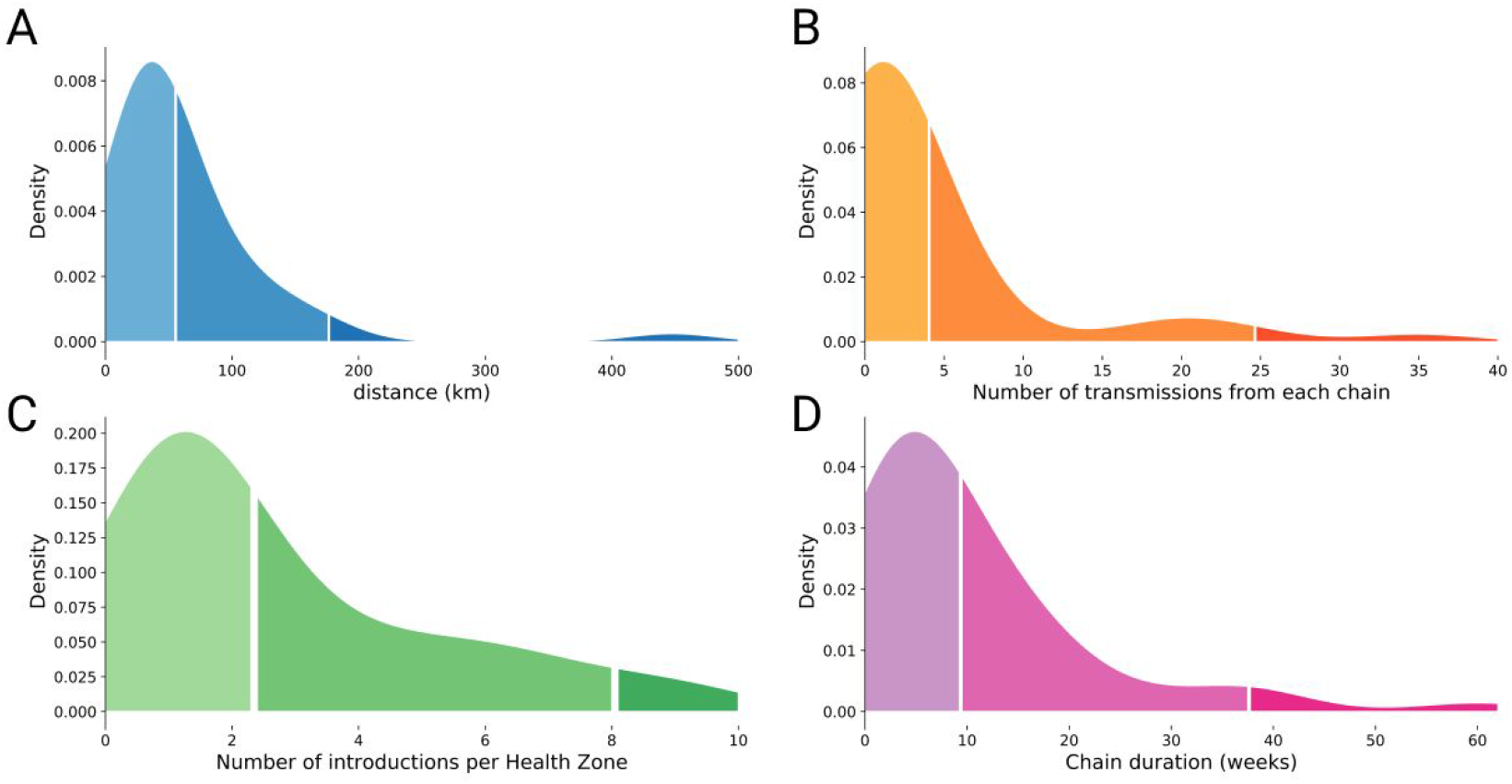
Transmission patterns within and between health zones. (A) Kernel density estimate of the inferred distance (in kilometers) between a source and a sink health zone, for 163 events where a transmission chain moved between two health zones; 50% of movement events occur between health zones that are less than 49km apart, and 95% of movement events occur between health zones that are less than 131km apart. (B) Kernel density estimate of the number of times a single chain circulating within a health zone seeded transmission into a different health zone; 50% of chains seed less than 1.5 transmissions into a different health zone, and 95% of transmission chains seed less than 6.2 transmissions into a different health zone. (C) Kernel density estimate of the number of times EBOV was introduced into each health zone; 50% of health zones experienced less than 7.1 introduction events, and 95% of health zones experienced less than 31.1 introduction events. (D) Kernel density estimate of the duration of time a transmission chain circulated within a single health zone; 50% of transmission chains circulated within a single health zone for less than 3.4 weeks, and 95% of transmission chains circulated within a single health zone for less than 20.9 weeks.

It can be useful to know how frequently EVD moves between different geographic areas. The policies instituted for controlling transmission when introductions and exportations are rare may be very different from the policies instituted when transmission chains move frequently between different regions. Using ancestral state reconstruction from the phylogeny, we assessed patterns of transmission chain movement across the entire outbreak. We detected 191 events where an EVD transmission chain moved from one health zone into a different health zone. Of these movement events, we were able to infer the source location with 70% certainty for 163 events, finding 57 distinct movement patterns of EVD between all possible pairwise combinations of affected health zones. Of the 26 affected health zones, 15 health zones acted only as sinks, meaning that once EVD transmission moved into them, they did not seed transmission into any further health zones (Supplemental Figure 2A). Fewer than half of health zones were identified as sources of transmission (*n =* 11/26), and the majority of EVD exportation events occurred from only 5 source health zones: Beni, Mabalako, Katwa, Kalunguta, and Mandima (Supplemental Figure 2A,C>). Each of these five health zones seeded transmission in a different health zone at least 20 separate times. Nineteen health zones received multiple introductions (Figure 2C, Supplemental Figure 2A). For each health zone, the number of introduction events into the health zone was correlated with the number of exportation events out of that health zone (r^2^=0.39, p<0.001, Supplemental Figure 2B).

In general, a circulating transmission chain was more likely to seed transmission in a different health zone if the two health zones were geographically close (Figure 2A), although we note that the geography and infrastructure of Eastern DRC means that straight-line distances may be misleading. The number of times that a transmission chain moved into a new health zone was variable (mean 3.7, standard deviation 6.8, Figure 2B). Just five transmission chains were responsible for 106 of the 163 events where a transmission chain moved into a new health zone (Supplemental Figure 2A,C). The duration of time that a chain circulated within a health zone was also variable (Figure 2D), and analysis of all transmission chains that moved between different health zones (*n* = 44) indicated that the length of time that a chain circulated in a health zone was weakly, but significantly, correlated with the number of times that transmission chain seeded transmission in other health zones (r^2^=0.19, p<0.003, Supplemental Figure 2D).

### Fine-scale transmission dynamics

For various infectious diseases, including seasonal influenza, Zika virus, and Ebola virus, genomic epidemiology studies have increased our understanding of transmission dynamics at broad spatiotemporal scales ^3,14–19^. While these large-scale descriptive inferences provide important context during outbreaks, frontline public health workers also need specific, actionable pieces of information. When responding to EVD outbreaks, analyses should provide information that supports contact tracing efforts and evaluates the effectiveness of surveillance systems and infection control measures ^20^. Here we describe the various types of fine-scale epidemiological dynamics that we have observed during the Nord Kivu EVD outbreak, and explain their importance to outbreak response efforts.

#### Sources and sinks of infections

Phylogeographic analyses, which infer patterns of transmission jointly from geographic and genomic data, reconstruct pathogen spatial movements over time^21^. From these reconstructed patterns we can describe which regions act as sources, that is they export transmission chains to other regions, and which regions act as sinks, meaning that they receive transmission chains but do not seed them elsewhere. Differentiating between source and sink regions can inform the allocation of surveillance resources. If a particular region is a primary source of transmission seeding transmission chains in other regions, then enhanced contact tracing and outreach efforts in the source area can limit transmission in both the source and sink regions.

Below we show an example of qualitative source and sink inference. Genomic data suggest that clade c26 circulated primarily in Mandima health zone (Figure 3, pink). Genomes from other health zones, including Butembo (clade c27, orange), Mambasa (c36 and c40, periwinkle), and Kayna (c28, teal), form their own clades interdigitating within clade c26. Under the assumption that sequenced cases are representative samples of EVD infections, this pattern suggests that Mandima acted as a source of the circulating chains, seeding transmission chains in Butembo, Mambasa, and Kayna that circulated within those health zones but did not move beyond them. These findings can be strengthened by analyzing case surveillance data for travel history or contact links across health zones.

**Figure 3:**
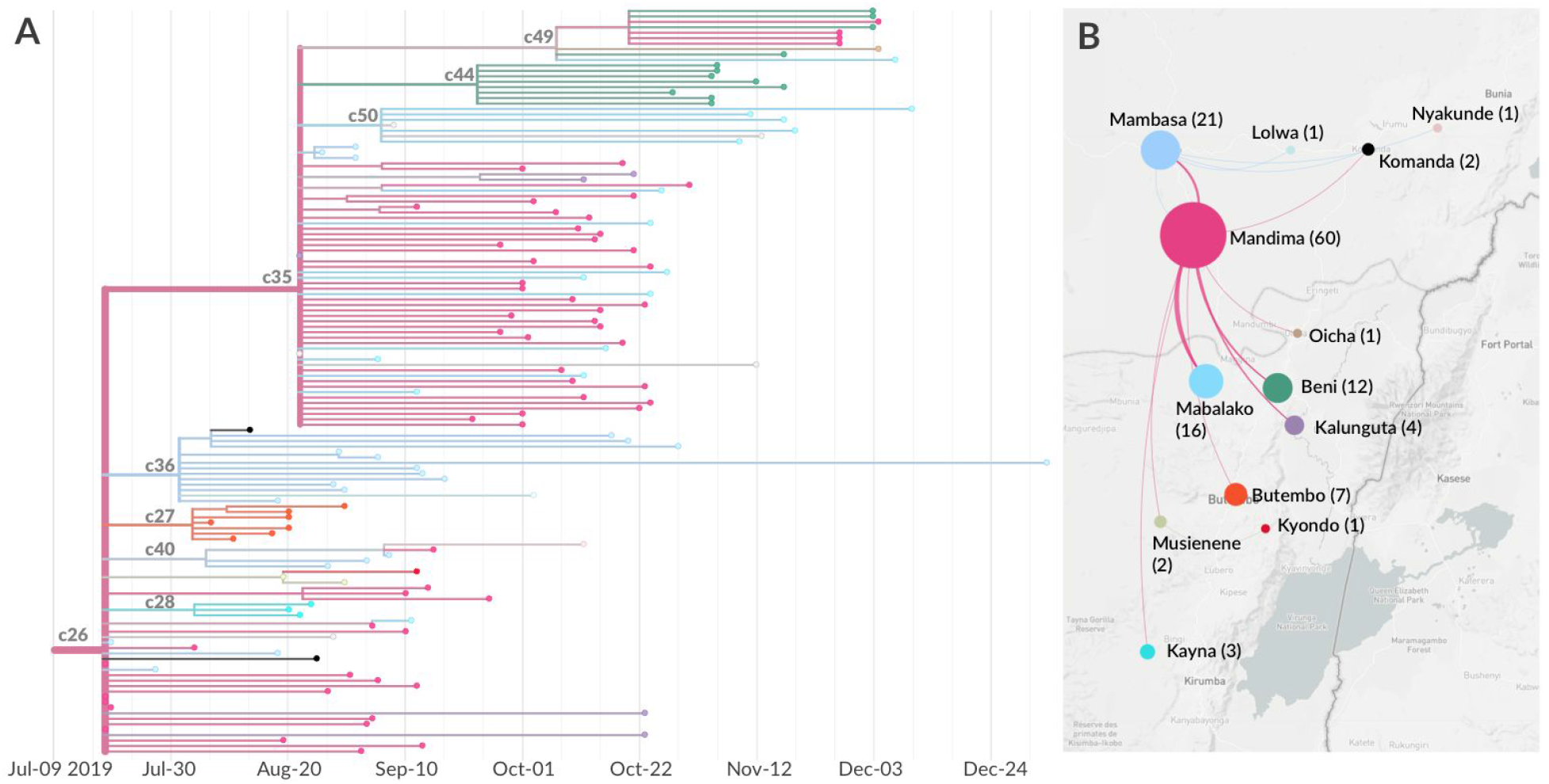
Source-sink transmission dynamics in Mandima health zone. (A) Phylogenetic reconstruction of a transmission chain circulating primarily in Mandima (c26), which moved into 12 other health zones; clades mentioned in the text are annotated on the phylogeny. (B) Geographic representation of the inferred transmission events between health zones. Circle size corresponds to sequenced sample counts for each health zone indicated. Counts are also annotated next to the health zone name.

#### Resolution of multiple concurrently circulating transmission chains

During large outbreaks, multiple separate transmission chains may circulate concurrently in the same geographic area ^3^. It may be difficult to differentiate between co-circulating transmission chains if exposure profiles are not distinct. Using molecular data increases the sensitivity with which we can define groups of related infections ^6^. Resolving distinct, co-circulating transmission chains is important because separate transmission chains may contribute disproportionately to overall case counts, or the factors that sustain transmission may vary by chain. Additionally, genomic data may detect transmission chains that are under-surveilled, something that can be masked in aggregate case count data. Thus defining clusters of related infections at high resolution may enable public health officials to evaluate and explicitly tailor surveillance and response measures.

In September 2019, we sequenced seven new cases from Kalunguta health zone (Figure 4A, enlarged tips); all had been collected between August 26th and September 1st, 2019. Despite coming from the same health zone and having similar sample collection dates, these cases were genetically diverse, clustering into three separate clades that each appeared to have resulted from a separate introduction from Beni (Figure 4A,B). These clades are c30, defined by a T to C mutation at nucleotide site 18083; c31, defined by a mutation from G to A at site 10982; and c32, defined by five mutations: T2695C, A3098G, C11678A, G11934A, and C13172T (Figure 4).

**Figure 4:**
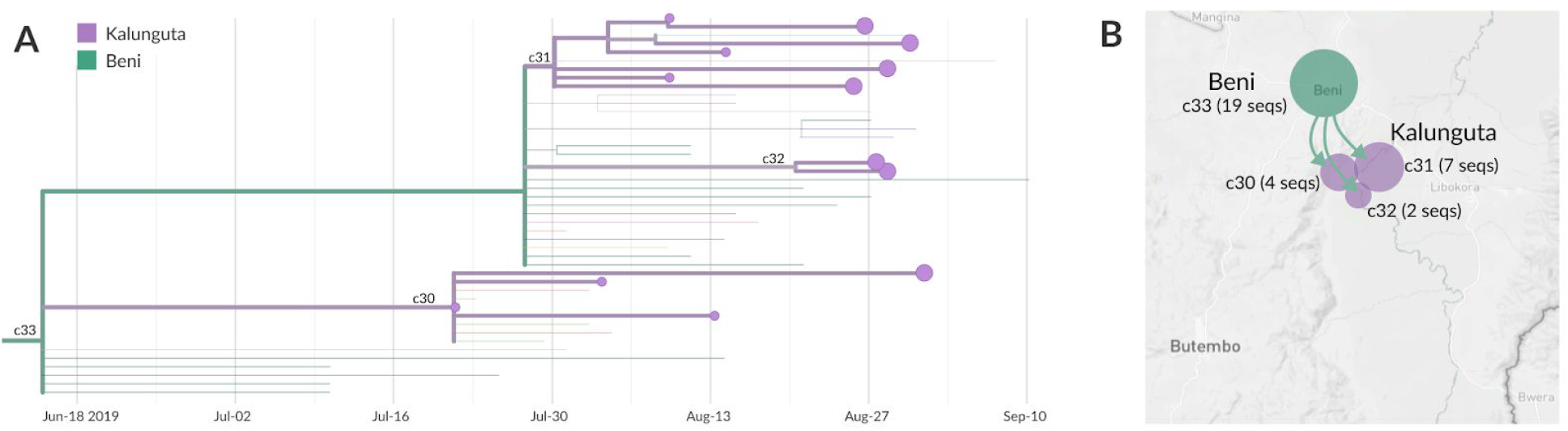
Multiple transmission chains co-circulating in Kalunguta health zone. (A) View of clade c33 and its descendents. Clade c33 circulated in Beni and was exported to Kalunguta three separate times, yielding three co-circulating transmission chains in Kalunguta (clades c30, c31, and c32). Enlarged tips show which samples were sequenced in September 2019, smaller tips were sequenced previously. Map schematic showing the inferred transmissions from Beni into Kalunguta for this clade; each paraphyletic introduction is represented by a labelled different circle, where the circle area is proportional to the number of sequences falling within the labelled clade. The exact count is also annotated.

Field epidemiologists hypothesized that the cases in c31 were part of a transmission cluster (data not shown to protect patient privacy). The genomic data reinforce this picture. In contrast, cases KAT10686 and BTB24923, which group together in clade c32, are significantly diverged from the other Kalunguta cases. This finding indicates that clade c32 was circulating in Kalunguta, during which time mutations accrued across the genome. However, this circulation was not detected by genomic surveillance until these two cases were sequenced. Given that we do not sequence all cases, this pattern indicates two possibilities. The first is that transmission was entirely cryptic, with neither genomic surveillance nor traditional surveillance capturing this chain of transmission within Kalunguta. If this were the case, the genomic data would indicate a need for enhanced contact tracing and community outreach within Kalunguta. Alternatively, this transmission chain may have been detected by traditional surveillance, but cases were simply not sent for sequencing, as may happen if reported contact histories are detailed and clear. This situation demonstrates the importance of having an integrated response team with expertise in interpretation of both genomic and case surveillance data; this integration would allow real-time differentiation of these two possible scenarios.

#### Superspreading

EVD can be transmitted during traditional burial practices, which may include frequent contact with the body of the deceased ^22^. Funerals for prominent figures in the community are often well attended, which can lead to superspreading events that cause large numbers of secondary cases and seed transmission in wider geographic areas ^23^. From a surveillance perspective, knowing that a superspreading event has occurred, and knowing which cases were infected as part of that event, can explain increased case counts in previously low incidence areas ^23^. Alternatively, if surveillance is enhanced in the wake of a known large exposure event, epidemiologists will likely detect prevalent cases unrelated to the superspreading event. These alternate exposure settings must also be investigated to control transmission. In both scenarios, genomic data enable classification of cases as related or unrelated to the superspreading event at a high resolution.

During this outbreak we have monitored the genomic data for possible superspreading events. We illustrate one such example, shown in clade c25. KAT5915 was a pastor who died of EVD in Beni. The body of the deceased was transported from Beni to Butembo for burial. Their funeral, which did not follow EVD safe burial protocols ^24^, was widely attended. While many people were exposed at the funeral, the genomic data indicate which infections were likely directly related to KAT5915. Exposure at the funeral led to further cases in Beni, and to the infection of individuals from Butembo, Ariwara, and Oicha (Figure 5). Three other cases had identical viral genome sequences to KAT5915, while another 5 cases had sequences that differed from KAT5915 by only one nucleotide (Figure 5A). This degree of sequence similarity indicates that infection likely occurred due to funerary contact. The genomic data also indicate that these funeral-associated infections started chains of transmission that continued to circulate and spread more widely. As of February 2020, 210 sequences across 20 health zones were descendents of this clade (Figure 5A,B).

**Figure 5:**
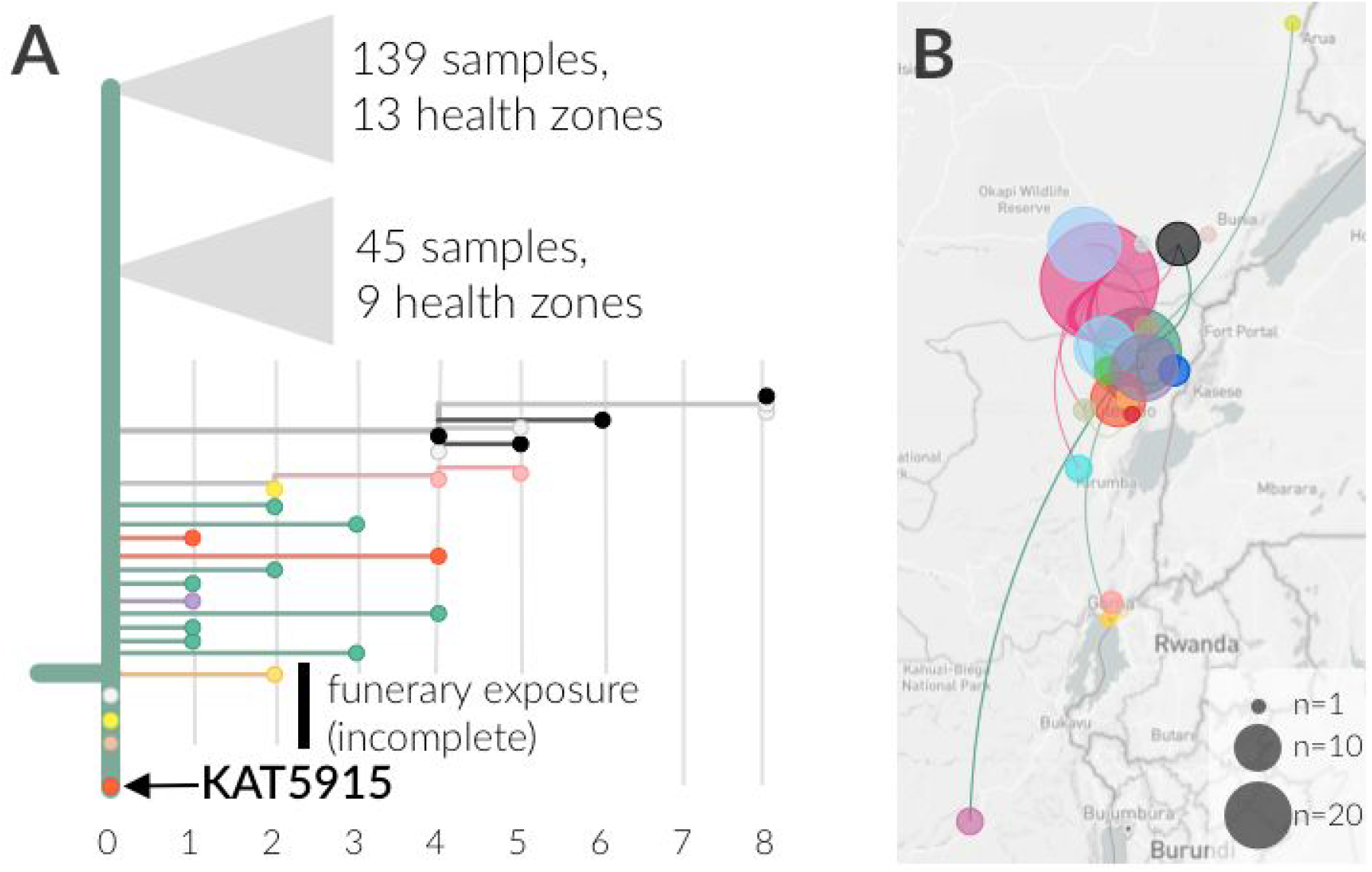
Superspreading event and sustained transmission after unsafe burial of a pastor. (A) Phylogenetic reconstruction of the funerary associated clade. The horizontal axis represents mutations relative to the index case (KAT5915, orange, labelled). Three other samples had identical genome sequences to KAT5915. One case was from Oicha (light brown), one case was from Ariwara (neon yellow), and one case lacked geographic information (grey). Additional cases diverged by only one nucleotide were detected in Beni (green), Butembo (orange), and Kalunguta (purple). In total, 210 sampled infections descend from this event, spanning 20 health zones. (B) Map indicating the numbers of sequences descended from this clade by health zone. The area of the circle is proportional to the number of sequenced cases from a health zone. Movements between health zones are shown as lines linking health zones.

#### Differentiating direct transmission from propagated transmission

Promptly detecting and isolating EVD cases can limit the risk of onward transmission to family and other contacts, lower the number of deaths that occur in the community, and potentially improve patient outcomes because care is sought earlier in the course of disease ^25^. Given the importance of finding all cases along a transmission chain, a critical component of EVD surveillance epidemiology is differentiating direct transmission events between two cases from longer transmission chains with intermediary infections. Genomic data are useful for detecting cryptic transmission, as viral genomes mutate during viral replication, leaving a history of infection even when a case is not captured by surveillance. This property allows us to assess the probability that a certain number of intermediate cases occurred between two sequenced cases given the number of nucleotide substitutions observed between their genomes.

We estimate the number of serial intervals along a transmission chain given observed genetic divergence using the following logic. EBOV has an evolutionary rate of roughly 1.2×10^−3^ substitutions per site per year ^26^. With a genome length of roughly 19,000 nucleotides, on average we expect 22.8 substitutions to accrue across the viral genome during a year of EBOV circulation. The EBOV serial interval, or the duration of time between symptom onsets in an index case and in a secondary case, is 14 to 15 days ^27^. Thus on average, 0.875 substitutions will accrue after a 14 day period. Notably these estimates are averages, but in the data one observes discrete numbers of substitutions. These substitutions accrue according to a Poisson process, and the probability of observing a specific number of mutations can be modeled as a Poisson distribution where the mean is the substitution rate per serial interval. In our case, given that two cases are directly linked and separated by a single 14 day serial interval, the probability of observing no substitutions is 0.42, the probability of observing one substitution is 0.36, and the probability of observing two mutations is 0.16. These probability distributions can be modeled for any number of serial intervals (Figure 6A).

**Figure 6:**
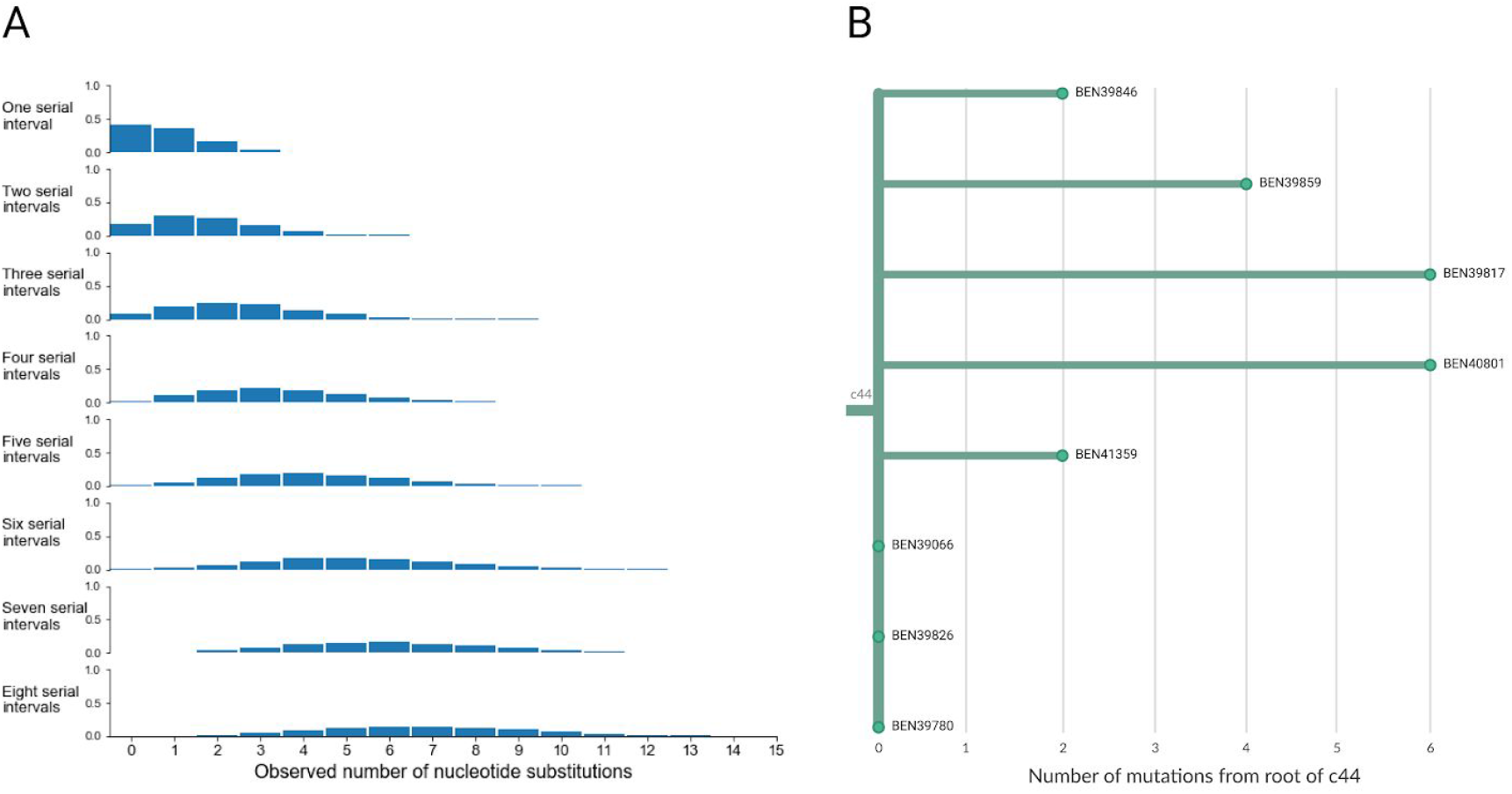
Expected numbers of mutations over transmission chains of different lengths, and application to transmission within Beni. (A) The distributions show the probability of observing a particular number of substitutions separating two sequences given that the two sampled cases are separated by a certain number of serial intervals. Here we model the probability distributions for one through eight serial intervals separating two cases. (B) Phylogenetic reconstruction of clade c44 showing genetic divergence between genomes. The branch lengths indicate the number of substitutions separating a sample from the root of clade c44. The three samples that are stacked vertically at the root of the clade, BEN39780, BEN39826, and BEN39066, all have identical consensus genome sequences.

As a concrete example we can look at clade c44, a cluster of 8 sequenced cases that circulated in Beni. These cases were sampled between October 28, 2019 and November 17, 2019, a period of 20 days. Three of these samples (BEN39066, BEN39826, and BEN39780) all have identical genome sequences (Figure 6B), which is consistent with either sequential direct transmission between the cases, or with a single source infecting multiple cases. In contrast, BEN39817, which was sampled on the same day as BEN39826 and BEN39780, is diverged from those sequences by 6 substitutions.The probability of observing 6 mutations separating two direct infections is negligible (*p* = 0.00026). The more probable explanation for this amount of genetic divergence is the presence of unsequenced cases along the transmission chain leading to the infection of BEN39817. As an additional note on interpretation, while both BEN39817 and BEN40801 are both diverged from the root of clade c44 by 6 mutations, BEN39817 has a *different* 6 mutations than BEN40801. That is, the mutations occurring along these branches are unique to that sample. This means that the intermediate infections prior to BEN39817’s infection are different from the intermediate infections leading to BEN40801.

## Discussion

In response to the ongoing Ebola outbreak in Nord Kivu, Democratic Republic of the Congo, we implemented an end-to-end genomic surveillance system. This system includes viral whole genome sequencing, bioinformatic analysis, and dissemination of genomic epidemiologic results to frontline public health workers. We used the genomic surveillance data to describe epidemic dynamics broadly, finding that while many health zones in Nord Kivu have been affected by EVD, a small number of transmission chains circulating within a limited subset of health zones have sustained the outbreak. These transmission chains moved frequently between this subset of health zones, likely increasing the challenge of containing the outbreak. We also used the genomic data to explore fine-scale dynamics that are actionable, such as precisely defining relatedness of infections, differentiating multiple co-circulating viral lineages, investigating superspreading events, and monitoring for possible undetected transmission chains. While these inferences have great public health utility, genomic epidemiologic training remains limited within the public health workforce. We therefore developed an innovative platform for communicating salient results, returning our interpretations of the genomic data within situation reports with narrative descriptions of epidemiologic dynamics accompanying genomic data visualizations.

In many ways, this outbreak shows how far genomic surveillance for outbreak response has come. At the time, the 2013-2016 West Africa EVD epidemic was notable for its high density of sequenced cases, representing around ∼5% of reported EVD cases ^3^. Within this outbreak, we have sequenced ∼17% of confirmed EVD cases, with all sequencing, and now most bioinformatic analysis, occurring within the DRC. The value of building capacity within-country is demonstrated not only by our work here, but also by the sustainability of a system that can be shifted to other surveillance efforts as well. Indeed, using this same genomic surveillance system we are now sequencing SARS-CoV-2 cases in the DRC.

While these are considerable strides, we have still encountered hurdles to generating data and communicating insights to the response team in a timely manner. We emphasize the critical importance of logistical support in the design of genomic surveillance systems. Although we have demonstrated that a one-week turnaround time between sample collection and return of results is possible, various obstacles have undermined our ability to always work that efficiently. Notably, challenges surrounding sample transport lengthened the average period of time between sample collection and generation of sequence data. This was especially true when periods of heightened violence caused the Katwa laboratory to close, and samples intended for sequencing in the field had to be shipped to Kinshasa, roughly 3000km away. Additionally, without a highly developed laboratory supply chain, it was hard to consistently access sequencing reagents, and lack of reagents sometimes delayed sequencing runs.

An additional consideration when performing genomic surveillance for outbreak response is how sampling may impact phylogeographic inference. Ideally, sequences should be sampled in proportion to incidence, and represent the full genetic diversity of the circulating pathogen. Genomic epidemiologic inferential techniques make these assumptions about samples, however we know that these assumptions are often violated by convenience sampling during outbreaks. Therefore, as genomic surveillance becomes more common, the field would benefit from additional simulation-based work exploring how genomic epidemiologic interpretations may change as a function of sampling. Finally, phylogenetic reconstructions may change with the addition of more sequence data. This does not necessarily mean that the reconstruction is wrong; rather, one can think of the reconstruction as incomplete due to lack of data. Increasing genomic surveillance capacity such that even higher proportions of cases can be sequenced will go far to alleviate these limitations.

The addition of genomic data to traditional epidemiologic data improves our ability to support contact tracing and evaluate interventions. Drawing inferences from multiple data sources can provide greater confidence in inferred epidemiologic dynamics, and also pinpoint weaknesses or erroneous findings in one data stream. Thus we emphasize the utility of reviewing findings from genomic surveillance and traditional surveillance in an integrated manner. Supporting integrated surveillance systems and epidemiologic response will require technical and social progress. From a technical perspective, we will need unified databases that provide all public health responders with access to well linked epidemiologic information, laboratory information, and genomic data for cases. From a social perspective, we envision that the utility of genomic surveillance will be improved if genomic and traditional epidemiologists work together, in real-time, during outbreak response. Performing rapid analysis with the opportunity for communication about the results enables an iterative discussion of the epidemiological dynamics within the context of both the genomic data and case history data. Facilitating this review process would increase the efficiency with which we can act on the results, leading to quicker evaluation and refinement of outbreak response efforts. While these next steps will require cooperation, openness, and motivation, we hope that our work shows the utility and worthiness of this effort.

## Data Availability

All genomic surveillance data, including consensus genomes and de-identified metadata, are publicly available at https://github.com/inrb-drc/ebola-nord-kivu. Nextstrain Augur and Auspice are open-source and all source code can be found at https://github.com/nextstrain/augur and https://github.com/nextstrain/auspice. All other analyses presented here are available as iPython notebooks at https://github.com/blab/ebola-narrative-ms/.

https://github.com/nextstrain/auspice

https://github.com/blab/ebola-narrative-ms/

## Acknowledgments

Sequencing activities were supported by the Defense Biological Product Assurance Office through a task order award to the National Strategic Research Institute (FA4600-12-D-9000) and Gates Foundation INV-004176 awarded to Catherine Pratt. AB was supported by the National Science Foundation Graduate Research Fellowship Program under Grant No. DGE-1256082. TB is a Pew Biomedical Scholar and is supported by NIH R35 GM119774-01. Computational infrastructure and in-country training was supported by the Fogarty International Center NIH/CRDF Global FOGX-19-90402-1 and the Bill and Melinda Gates Foundation INV-003565. The content of this Article does not necessarily represent the official policy or views of the US Department of the Army, the US Department of Defense, the US Department of Health and Human Services, the US Government, or the institutions or companies affiliated with the authors.

